# An approach for early diagnosis of atherosclerosis: correlation between blood levels of endothelin and D-dimers with atherogenic index in African black subjects

**DOI:** 10.1101/2024.12.27.24319552

**Authors:** Claudric Roosvelt Tchame, Euloge Yiagnigni Mfopou

## Abstract

Global mortality due to cardiovascular disease (CVDs) is continuously increasing. In this regard, several authors have investigated the origins of CVD, with atherosclerosis being the most involved pathological process in CVD. In our study, we aimed to measure some parameters (endothelin, D-dimers) that could be involved in the early diagnosis of atherosclerosis and compare them to a classical marker Atherogenic index (AI) to see if there would be a correlation between these parameters and a possible use of them in diagnosis.

To do this, patients at risk of atherosclerosis, hypertensive and diabetic, were recruited over a period of 5 months at the Bonne Santé clinic in Yaounde. Out of a population of 84 patients, 10 hypertensive diabetics were selected, 77 hypertensive and 17 diabetics. The data analysis was conducted using the Spearman’s Rho test, after which we did not observe any significant correlation between endothelin and AI, with a correlation coefficient of 0.065 and a P value of 0.556. However, a significant correlation between D-dimers and AI was observed, with a correlation coefficient of 0.231 and a P value of 0.034; and a significant negative correlation between endothelin and D-dimers was observed, with a correlation coefficient of -0.232 and a P value of 0.033.

By comparing these data with those from other articles, we concluded that endothelin alone is not a good diagnostic marker for atherosclerosis. However, D-dimers can be used as associated markers for atherosclerosis diagnosis. These results directed us towards the process and different initiatic factors of atherosclerosis.

## 1. Introduction

Cardiovascular diseases (CVDs) are now the leading cause of mortality worldwide (1). WHO estimates that 17.7 million deaths are attributable to cardiovascular diseases, accounting for 31% of total global mortality (1). Non-communicable diseases such as cardiovascular diseases and diabetes are increasingly becoming the leading cause of mortality in Sub-Saharan Africa, where they accounted for 37% of mortality in 2019, compared to 24% in 2000 (2). In Africa, the increasing prevalence of CVDs has become a major public health concern. CVDs can be associated with other pathologies such as arterial hypertension and diabetes. Arterial hypertension remains a concern in all African countries, where it accounts for 20 to 30% of hospital admissions (3). In a Sub-Saharan African country, specifically Cameroon, the prevalence of simple arterial hypertension is approximately 30%, while severe arterial hypertension is approximately 6% (4). Cardiovascular diseases (CVDs) represent the most significant cause of morbidity and mortality among patients with type 2 diabetes.(5) While the number of people living with diabetes is expected to reach 47 million by 2045, compared to 19 million in 2019 (2). Ischemic heart diseases are the leading cause of mortality worldwide, and atherosclerosis is therefore the pathological process most involved in global mortality (6). Following this information, we aimed to recruit patients at high risk of developing atherosclerosis, namely hypertensive and diabetic patients, for whom we measured several parameters including type 1 endothelin (ET 1), which is mainly found in the vascular endothelium, D-dimers, which are fibrin degradation products used for the diagnosis of thromboembolic diseases, and Total Cholesterol and HDL for atherogenic index (AI) calculation. We attempted to verify the hypothesis that endothelin found in the vascular endothelium could increase in the blood during early atherosclerotic lesion and be measured to indicate early atherosclerotic processes.

The classical methods for evaluating atherogenic risk consisted of evaluating lipoprotein levels through the atherogenic index (Total Cholesterol / HDL). Nowadays, there are several diagnostic techniques for atherosclerosis, but the problem still remains regarding early diagnosis and accessibility for the general public. To address this issue of early diagnosis of cardiovascular diseases that could be used for the general public, we decided to study various possible correlations between AI and two other parameters, ET 1 and D-dimers. These two markers utilized in our study were Endothelin Type 1 and D-dimers. Thus, we attempted to verify if an increase in blood ET 1 could be correlated with an increase in AI, or if an increase in D-dimers could be correlated with an increase in AI to demonstrate a similarly pathological process between the parameters. Our overall objective was therefore to determine the levels of endothelin and D-dimers in a high-risk population for atherosclerosis (hypertensive and diabetic patients) in comparison to the atherogenic index.

## 2. METHODOLOGY

### 2.1. Study desigh

We conducted a cross-sectional and analytical study from July 20th to November 24th, 2020, at the Clinique les Promoteurs de la bonne santé. This clinic is located in Yaoundé and offers several services, including a cardiology department and a diabetology department. To conduct this study, we selected patients at risk of developing established atherosclerosis processes.

We included hypertensive and diabetic subjects recruited from the Clinic in our sample. To ensure better quality of our results, all individuals with a pathology or physiological state that could influence the interpretation of the results were not included, such as those with a hematoma, smokers, and pregnant women.

Throughout the course of our study, we obtained written and verbal consent from all participants, with the minimum age of our population being 39 years old. Following the patient’s agreement to participate, we administered a comprehensive questionnaire designed to collect information on age, sex, medical history pertaining to diabetes or hypertension, as well as current medications consumed by the patient. Subsequently, samples were collected from each of the participants per the protocol.

### 2.2. Laboratory analysis

Samples were taken from the selected patients in 02 tubes, a dry tube for the analysis of the lipid profile (total cholesterol and HDL) and the analysis of Endothelin Type 1 (ET 1), and a citrated tube for the analysis of D-dimers. Specified equipment was used for each analysis, thus for the lipid profile, we used a spectrophotometer, an ELYSA chain for the dosage of ET 1, and an AFIAS automate for the dosage of D-dimers. The different equipment was pre-set in accordance with the analysis technical specifications provided by each manufacturer. The analyses were conducted per day, except for ET 1, where samples needed to be run in batches. Therefore, after analyzing other tests, the remaining serum was aliquoted and stored at -20°C for 2 weeks, and analyzed in 2 batches.

The laboratory analyses were performed as follows:

- For Endothelin (ET 1), we opted for the ELISA method using the Human Endothelin 1 (ET-1) ELISA Kit from RAYBIOTECK.
- For D-dimers, we used a sandwich immunodetection method with the AFIAS D-dimers instrument.
- For Total Cholesterol and HDL, we used a biochemistry assay by spectrophotometry, using the Biolabo manufacturer’s reagents.

### 2.3. Study population

The study population was calculated using the formula N = **(t² × p × (1-p)) / m²**, where:

- N: Minimum sample size required for obtaining significant results for a given event and a fixed level of risk.
- t: Confidence level (the standard value for 95% confidence level is 1.96).
- p: Estimated proportion of the population (hospitalized) that presents the characteristic, in this case, hypertensive patients at 10% in Cameroon.
- m: Margin of error (usually set at 5%).

By applying this formula, we obtained:

N = (1.96)² * 0.0644 (1-0.0644) / (0.05)²
N = 92 patients.

During our study, the size of our sample at the end of the study was N = **84 patients.**

### 2.4. Statistical analyses

The different data of each patient were recorded in CsPro 7.3 statistical software and analyzed using SPSS. The results were presented in the form of tables and figures. The correlations between D-dimers and atherogenic index, between endothelin and atherogenic index, and between endothelin and D-dimers were determined. Depending on the population distribution, we used the Spearman’s Rho test.

### 2.5. Ethical consideration

#### 2.5.1. Ethical approval number and research authorization

2.5.1. Ethical approval number and research authorization.
| Name | Ethical committee | Number | Method |
| --- | --- | --- | --- |
| TCHAME Claudric<br>Roosvelt | Université des<br>montagnes, bangangté | 71 | Analytical cross-sectional<br>study. |

##### 2.5.1.1. Research authorization

Conducted at the “Clinique des promoteurs de la bonne santé” in yaounde Cameroon, the research authorization was issued by Professor EULOGE YIAGNIGNI, who is the director of the clinic and co-author of the article.

#### 2.5.2. Informational letter

##### Participant Information Document (PID)

**Title of the Project:** –Early Diagnosis of Atherosclerosis by Assessing Endothelin and D-Dimers Level Based on the Atherogenic Index in Hospitals in Yaounde–

**Name of Site Investigator:** TCHAME Claudric Roosvelt

**Name of the Institute:** Mountains University. “Université des montagnes, bangangté”

**Objective:** Our main objective is to determine whether endothelin and D-dimers can be used as an early diagnostic tool.

**Protocol:** Cardiovascular diseases are currently the leading cause of death worldwide, often resulting from atherosclerotic lesions, particularly in the coronaries and cerebral circulation. In order to explore atherosclerosis, we plan to measure endothelin and D-dimers levels in association with the atherogenic index. Patients with hypertension or diabetes will be selected, Examinations will be carried out on the patients, including endothelin, D-dimer, total cholesterol, and HDL. Finally, different correlations will be determined between these parameters within our population.

The Ethics Committee of the Mountains University approved this protocol.

**Risks and Discomforts:** There are no potential risks associated with the sampling.

**Benefits and Financial Considerations:** Participation in the study will not cost you anything. A better understanding of the atherosclerosis initiation process will improve the onset of cardiovascular diseases and prevent their recurrence. Participants will have their examination results free.

**Confidentiality:** The participating sites and personnel will strictly respect your anonymity throughout the study and when communicating the results.

**Right to Refuse:** You are entirely free to accept or refuse participation in this study without any prejudice.

**Contacts:** For further information, please contact the researcher and investigator: TCHAME Claudric Roosvelt, Phone: 698104069 / 670751356.

#### 2.5.3. CONSENT FORM

Researcher’s commitment

Me, TCHAME Claudric Roosvelt commits me to conduct this study in accordance with all the ethical standards that apply to projects involving the participation of human subjects.

Participant’s consent

I, ____ [participant name] ____, confirm that I have read and understood the project information leaflet on, endothelin and dimer assay in the early diagnosis of atherosclerosis

. I have understood the conditions, the risks and the possible benefits of my participation. All my questions have been answered to my satisfaction. I had ample time to reflect on my decision to participate or not. I understand that my participation is entirely voluntary and that I may withdraw at any time without prejudice.

|  |  |
| --- | --- |
|  | Contact of researcher: |
| Date and Signature of Participant: | Date and signature of researcher: |

## 3. Result

### 3.1. Population profile

Our study population consisted of 84 patients, including 32 men and 52 women, with a male-to-female ratio of 0.61. The mean age was 62 years, with a minimum of 39 years and a maximum of 84 years. The mode of our statistical series was made up of people aged between 55 and 64 years. There were 77 hypertensive patients in our population, including 27 men and 50 women. There were 17 diabetic patients, including 10 men and 7 women. In our population, we had 10 patients who were both hypertensive and diabetic. Regarding medication intake, the average number of medications consumed in the population was 2 per individual, with a maximum of 5. None of our patients were taking statins, and the main medication used for the treatment of diabetes was metformin.

In our study population, all hypertensive and diabetic patients were on medication therapy except for about 3 patients, two of whom had just been diagnosed with hypertension, and one who was in the early stages of diabetes diagnosis. The results that follow will present laboratory data analyses.:

**Table 1:**
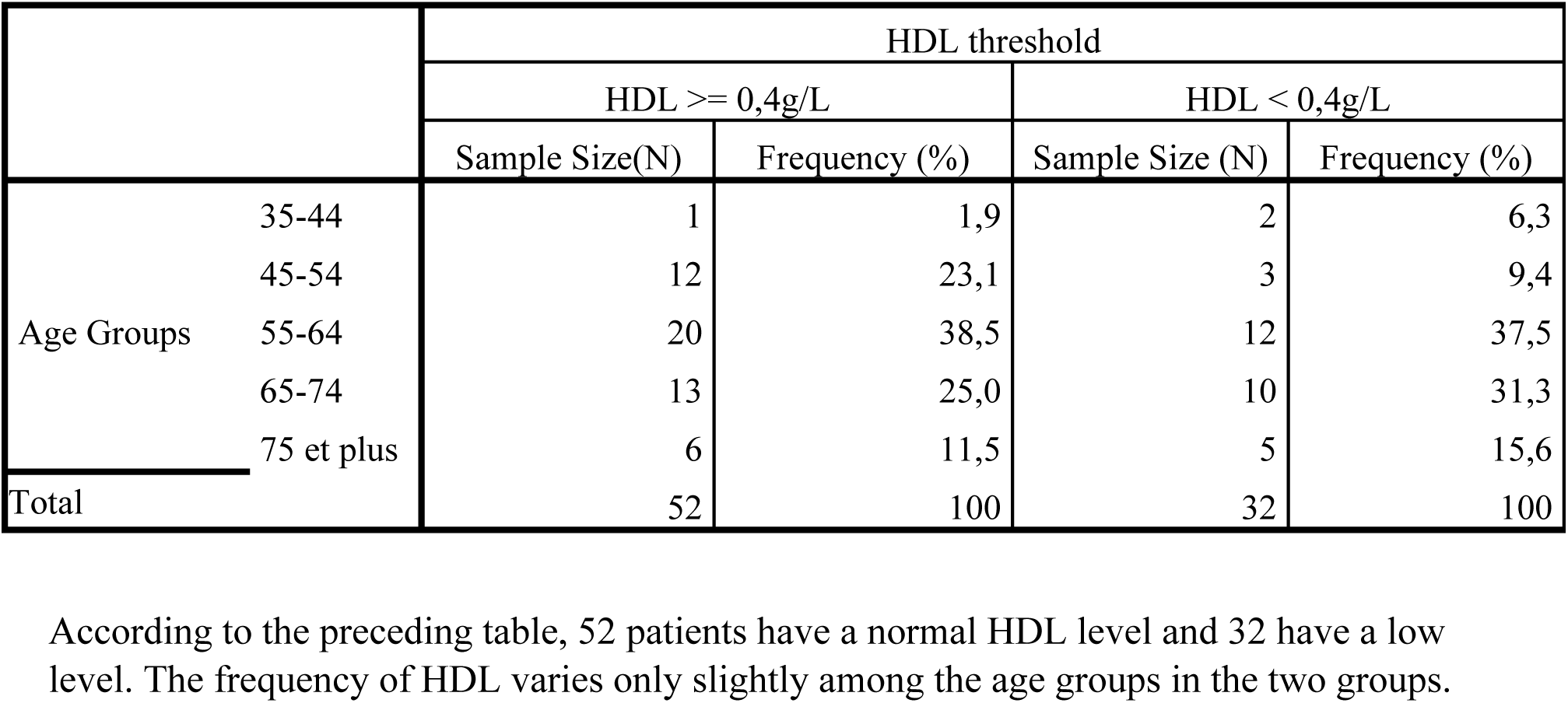
Distribution of the study population based on HDL threshold values according to age groups.

**Table 2:** Distribution of the study population based on total cholesterol threshold values according to age groups.

|  |  | ChT threshold |  |  |  |
| --- | --- | --- | --- | --- | --- |
|  |  | <2g/l |  | >=2g/l |  |
|  |  | Sample Size (N) | frequency (%) | Sample Size (N) | Frequency (%) |
| Age Groups | 35-44 | 3 | 4,92 | 0 | 0,0 |
|  | 45-54 | 8 | 13,11 | 7 | 30,4 |
|  | 55-64 | 22 | 36,07 | 10 | 43,5 |
|  | 65-74 | 19 | 31,15 | 4 | 17,4 |
|  | 75 et plus | 9 | 14,75 | 2 | 8,7 |
| Total |  | 61 | 100 | 23 | 100 |

According to the preceding table, 61 patients have a normal total cholesterol level, while 23 have a high level. The frequency of total cholesterol varies among the age groups in both groups.

**Table 3:** Distribution of the study population based on atherogenic index threshold values according to age groups.

|  |  | IA threshold |  |  |  |
| --- | --- | --- | --- | --- | --- |
|  |  | IA < 5 |  | IA > = 5 |  |
|  |  | Sample Size (N) | Fréquence (%) | Sample Size (N) | Frequency (%) |
| Age Groups | 35-44 | 3 | 4,6 | 0 | 0,0 |
|  | 45-54 | 12 | 18,5 | 3 | 15,8 |
|  | 55-64 | 26 | 40,0 | 6 | 31,6 |
|  | 65-74 | 16 | 24,6 | 7 | 36,8 |
|  | 75 et plus | 8 | 12,3 | 3 | 15,8 |
| Total |  | 65 | 100 | 19 | 100 |

**Table 4:**
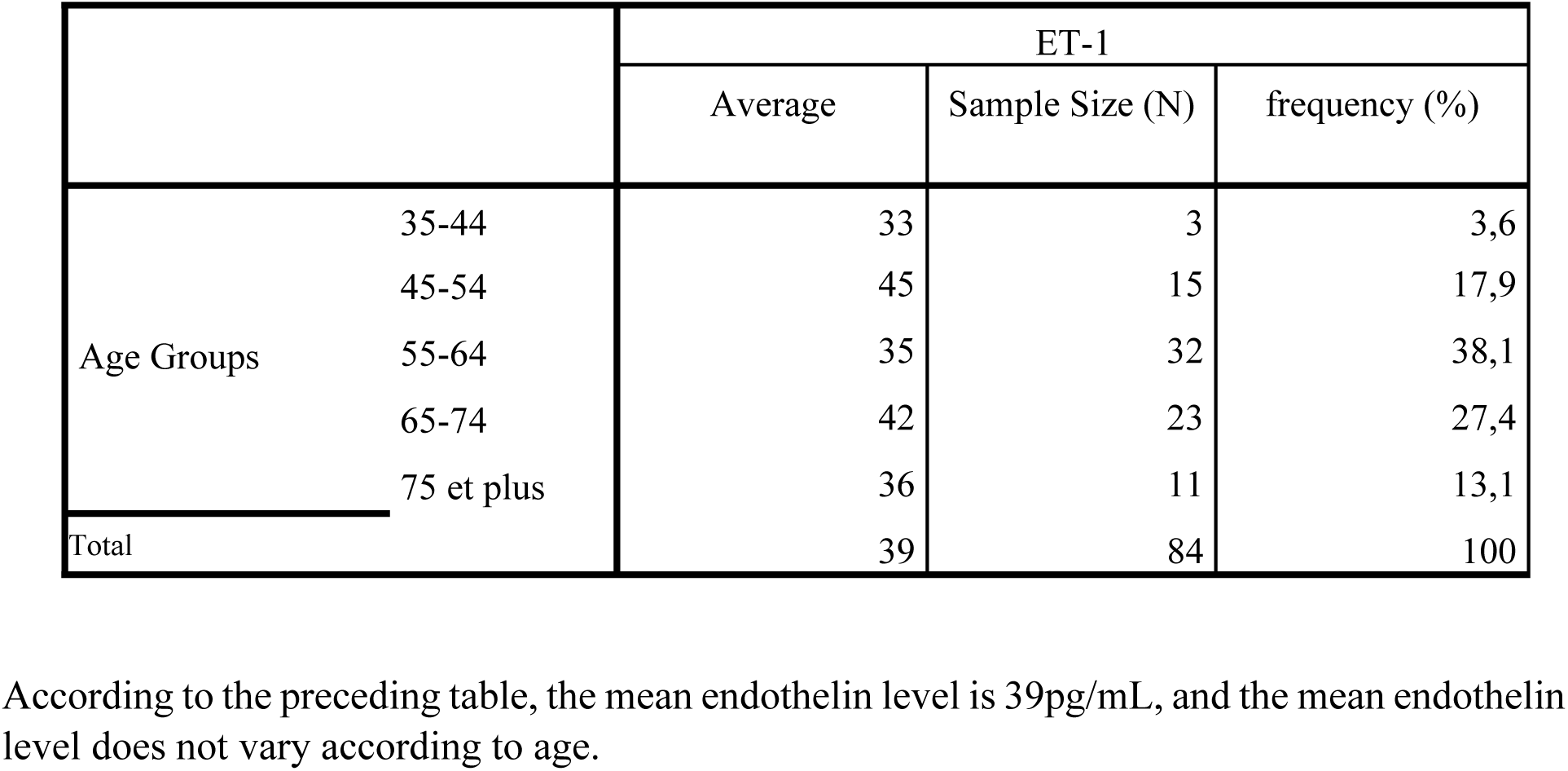
Variation of endothelin means according to age groups.

|  |  | ET-1 |  |  |
| --- | --- | --- | --- | --- |
|  |  | Average | Sample Size (N) | frequency (%) |
| Age Groups | 35-44 | 33 | 3 | 3,6 |
|  | 45-54 | 45 | 15 | 17,9 |
|  | 55-64 | 35 | 32 | 38,1 |
|  | 65-74 | 42 | 23 | 27,4 |
|  | 75 et plus | 36 | 11 | 13,1 |
| Total |  | 39 | 84 | 100 |

According to the preceding table, the mean endothelin level is 39pg/mL, and the mean endothelin level does not vary according to age.

**Table 5:** Variation of D-dimers according to age groups.

|  | D-Dimer threshold |  |  |  |  |  |  |  |
| --- | --- | --- | --- | --- | --- | --- | --- | --- |
|  | [0, 500[ |  | [500, 1000[ |  | [1000, 2000[ |  | 2000 et plus |  |
|  | Sample Size | Frequency (%) | Sample Size | Frequency (%) | Sample Size | Frequency (%) | Sample Size | Frequency (%) |
| 35-44 | 2 | 4,0% | 0 | 0,0% | 0 | 0,0% | 1 | 12,5% |
| 45-54 | 14 | 28,0% | 0 | 0,0% | 0 | 0,0% | 1 | 12,5% |
| 55-64 | 19 | 38,0% | 7 | 41,2% | 3 | 33,3% | 3 | 37,5% |
| 65-74 | 10 | 20,0% | 10 | 58,8% | 2 | 22,2% | 1 | 12,5% |
| 75 et plus | 5 | 10,0% | 0 | 0,0% | 4 | 44,4% | 2 | 25,0% |
| Total | 50 | 100% | 17 | 100% | 9 | 100% | 8 | 100% |

### 3.2. Analysis of different linear regressions

Two correlation tests were carried out, first between endothelin and D-dimers based on total cholesterol and HDL, and then between endothelin and D-dimers based on the atherogenic index.

The following table shows the different correlations that exist between endothelin, D-dimers, total cholesterol, and HDL:

| Corrélations |  |  | ET 1 | ChT | D-Dimer | HDL |
| --- | --- | --- | --- | --- | --- | --- |
| Rho de Spearman | ET 1 | Correlation Coefficient | 1,000 | ,281** | -,232* | ,139 |
|  |  | P-value | . | ,010 | ,033 | ,208 |
|  |  | N | 84 | 84 | 84 | 84 |
|  | ChT | Correlation Coefficient | 0,281** | 1,000 | -,205 | 0,358** |
|  |  | P-value | 0,010 | . | ,061 | 0,001 |
|  |  | N | 84 | 84 | 84 | 84 |
|  | D-Dimer | Correlation Coefficient | -,232* | -,205 | 1,000 | -,392** |
|  |  | P-value | ,033 | ,061 | . | ,000 |
|  |  | N | 84 | 84 | 84 | 84 |
|  | HDL | Correlation Coefficient | ,139 | ,358** | -,392** | 1,000 |
|  |  | P-value | ,208 | ,001 | ,000 | . |
|  |  | N | 84 | 84 | 84 | 84 |

According to the analysis of Spearman's rho, there is a significant correlation between ET-1 and total cholesterol, with a p-value of **0.010** and a correlation coefficient of **0.281**. There is a significant correlation between HDL and total cholesterol, with a p-value of **0.001** and a correlation coefficient of **0.358**.

The following table shows the different correlations that exist between endothelin and atherogenic index, D-dimers and atherogenic index, and finally between endothelin and D-dimers.

**Tableau 7:** Different correlation tests.

| Corrélations |  |  | EDN1 | D-Dimères | IA |
| --- | --- | --- | --- | --- | --- |
| Rho de Spearman | EDN1 | Correlation Coefficient | 1,000 | -,232* | ,065 |
|  |  | P-value | . | ,033 | ,556 |
|  |  | N | 84 | 84 | 84 |
|  | D-Dimère | Correlation Coefficient | **-0,232\*** | 1,000 | ,231* |
|  |  | P-value | **0,033** | . | ,034 |
|  |  | N | 84 | 84 | 84 |
|  | IA | Correlation Coefficient | **0,065** | **0,231\*** | 1,000 |
|  |  | P-value | **0,556** | **0,034** | . |
|  |  | N | 84 | 84 | 84 |

According to Spearman's rho analysis, there is no significant correlation between endothelin and atherogenic index, as the correlation coefficient is very low (**0.065**) and the p-value is above 0.05 (**0.556**).
There is a significant correlation between D-dimers and atherogenic index, as the p-value is **0.034** for a correlation coefficient of **0.231**.
There is a significant negative correlation (p-value = **0.033**) between EDN1 and D-dimers, and their Spearman correlation coefficient is **-0.232**.

## 4. Discussion

In our study population, we observed a strong and significant correlation between HDL and total cholesterol, with a p-value of 0.001 and a correlation coefficient of 0.358. Total cholesterol is mostly comprised of LDL, which transports cholesterol to tissues, while HDL retrieves excess cholesterol from vessels and metabolizes it in the liver. Therefore, we should observe a simultaneous increase in these two parameters, which was the case in our study, indicating the quality of our data. It should be noted that in our population, 65 out of 84 patients had a normal atherogenic index.

After analyzing Spearman’s rho, a significant correlation between ET-1 and total cholesterol with a p-value of 0.010 and a correlation coefficient of 0.281 was observed in our population, which is consistent with the results of several studies. For example, a study conducted in 1993 by Lerman A, et al showed that pigs fed a high cholesterol diet had an increase in plasma ET-1 levels (7). In 2006, Jianglin Fan, et al made a similar observation: endothelin (ET) receptors are upregulated in human and laboratory animals with atherosclerotic lesions. Later, they tried to find the role of HDL and LDL in endothelin (ET) secretion and were able to show that both normal LDL (nLDL) and oxidized LDL (oxLDL) can act as potential endogenous mediators of ET-1 release in atherosclerosis, while HDL may exert an anti-atherogenic action by inhibiting the polarized release of ET-1 (8).

After analyzing the Spearman correlation, no significant correlation between ET-1 and AI was observed. However, the expression and immunoreactivity of ET-1 are upregulated in atherosclerosis (9). The total cholesterol/HDL ratio and the LDL/HDL ratio, which are atherogenic indices, are reliable markers for assessing the risk of developing a cardiovascular event (10), Our results suggest that endothelin alone cannot be used to diagnose atherosclerosis. The increase in endothelin is not necessarily pathological in the body, as several stimuli can induce its production (11). Indeed, when using a protein in a diagnostic, it is essential to ensure its specificity and sensitivity with respect to the disease. This evaluation of specificity and sensitivity is done using a reference test. However, regarding cardiovascular disease, there is no reference test that can accurately determine the presence of the disease due to its metabolic nature. We only have markers indicating an increased risk of developing the disease. This is one of the reasons why we opted for this methodological approach of comparing certain new markers to other older ones already used in routine diagnosis. Faced with our results, we wondered about the process of initiating atherosclerosis. In an issue of the American Journal of Hypertension by Zhang et al., exploration of the interaction between ET-1, endothelial cells and macrophages in the context of atherosclerosis was conducted (12). They first made two observations; firstly, apolipoprotein-E-deficient mice pre-fed with a high-fat diet had an increase in ET-1 protein expression from endothelial cells in atherosclerotic plaques; secondly, endothelial cells from human umbilical veins demonstrated an increase in ET-1 production when exposed to oxidized LDL (oxLDL) (13). Based on these findings, the authors hypothesized that oxLDL stimulates endothelial ET-1 production. Thus, they show that a combination of ET-1 overexpression and oxLDL promotes endothelial cell production of a range of adhesion molecules and chemokines. These effects are not observed only in the case of excess ET-1, but a combination of ET-1 and oxLDL shows greater effects than those observed with oxLDL alone. This discovery led the same group to another study that ensured that ET-1 alone has no effect; a combination of ET-1 and oxLDL promotes macrophage migration. When macrophages were cultured with a conditioned medium from HUVECs (Human Umbilical Vein Endothelial Cells) treated with oxLDL, they developed more of an inflammatory M1 phenotype and less of an anti-inflammatory M2 phenotype, as demonstrated by up-regulation of interleukin encoding messages (13). These different observations, combined with our results, allow us to confirm that several conditions must be met for the initiation of an atherosclerotic process and to reject the isolated use of endothelin for the diagnosis of cardiovascular diseases.

In our study, a significant correlation was observed between (D-DIMERS, AI) with a correlation coefficient of 0.231 and a P value of 0.034. D-dimers are markers of fibrinolysis that specifically involve fibrin clots. Normal or low levels of D-dimers can rule out suspicion of a thromboembolic process. Baseline levels of D-dimers can be used as a biomarker for assessing the risk of stroke (14). Thus, Kang-Ho Choi, et al demonstrated through a completed cohort in 2012 and published in 2016 that a high baseline concentration of D-dimers can be used as a risk factor for ischemic stroke (15). We can say that D-dimers may have a real utility in the diagnosis of atherosclerosis.

In case of endothelial dysfunction, there is a release of endothelin and thromboxane A2 which have vasoconstrictive properties (16). Thromboxane A2 is a powerful inducer of platelet aggregation during plasma coagulation. D-dimers explore fibrinolysis, a process that involves the dissolution of fibrin clots to maintain blood in a fluid state in blood vessels after coagulation and thus promotes vessel repair. In normal physiological situations, after vessel coagulation and repair, the levels of ET-1 and thromboxane A2 should decrease, giving way to fibrinolysis, which was verified in our study. A significant negative correlation was observed between (ET-1, D-DIMER) with a correlation coefficient of -0.232 and a P value of 0.033. Thus, in the body, when ET-1 increases, D-dimers tend to decrease or when D-dimers increase, ET-1 tends to decrease. A simultaneous increase in D-dimers and endothelin would therefore suggest a pathological process that is not necessarily atherosclerotic. A study conducted in 2018 by Ronghua Gao, et al showed the importance of combining levels of D-dimer and ET-1 as predictive and prognostic values for acute ST-segment elevation myocardial infarction (17).

## 5. Conclusion

In our study, we observed that ET-1 could play a key role in the development of atherosclerosis, not only as a pro-inflammatory factor but also by influencing the behavior of immune cells within the atherosclerotic plaque. However, the precise mechanisms by which ET-1 contributes to the pathogenesis of atherosclerosis are yet to be elucidated. It is possible that ET-1 acts in synergy with other factors such as inflammatory cytokines and oxidative stress to promote disease initiation and progression. Although we established a significant correlation between ET-1 levels and total cholesterol, indicating an increase in endothelin during lipid disorders, ET-1 cannot be considered in isolation as a reliable diagnostic marker for cardiovascular diseases. Our study revealed that an association between Endothelin and oxLDL is more representative of an atherosclerotic pathology and that the association between D-dimers and ET-1 could be indicative of a pathological process, particularly in the chronic phases of the disease, where thrombosis occurs later.

A more in-depth analysis of the mechanisms by which ET-1 and other biomarkers interact and contribute to atherosclerosis is necessary. A better understanding of these interactions will enable us to develop more precise biological assessment methods for cardiovascular risk, taking into account the kinetics of the appearance of different markers depending on the stage of the disease. This could have significant implications for public health, allowing for better prevention and improved management of cardiovascular diseases.

## Data Availability

All data produced in the present work are contained in the manuscript

